# Prevalence and severity of symptoms 3 months after infection with SARS-CoV-2 compared to test-negative and population controls in the Netherlands

**DOI:** 10.1101/2022.06.15.22276439

**Authors:** Tessa van der Maaden, Elizabeth N. Mutubuki, Siméon de Bruijn, Ka Yin Leung, Hans Knoop, Jaap Slootweg, Anna D. Tulen, Albert Wong, Albert Jan van Hoek, Eelco Franz, Cees C. Van den Wijngaard

## Abstract

**Background:** More information is needed on prevalence of long-term symptoms after SARS-CoV-2-infection. This prospective study assesses symptoms three months after SARS-CoV-2-infection compared to test-negative and population controls, and the effect of vaccination prior to infection.

**Methods:** Participants enrolled after a positive (cases) or negative (test-negative controls) SARS-CoV-2-test, or after invitation from the general population (population controls). After three months, participants indicated presence of 41 symptoms, and severity of four symptoms. Permutation tests were used to select symptoms significantly elevated in cases compared to controls and to compare symptoms between cases that were vaccinated or unvaccinated prior to infection.

**Findings:** Between May 19^th^ and December 13^th^ 2021 9166 cases, 1698 symptomatic but test-negative controls, and 3708 population controls enrolled. At three months, 13 symptoms, and severity of fatigue, cognitive impairment and dyspnoea, were significantly elevated between cases and controls. Of cases, 48.5% reported ≥1 significantly elevated symptom, compared to 29.8% of test-negative controls and 26.0% of population controls. Effect of vaccination could only be determined for cases <65yrs, and was found to be significantly protective for loss of smell and taste but not for other symptoms.

**Interpretation:** Three months after SARS-CoV-2 infection, almost half of the cases still report symptoms, which is higher than the background prevalence and prevalence in test-negative controls. Vaccination prior to infection was protective against loss of smell and taste as assessed in cases aged <65.

## INTRODUCTION

The SARS-CoV-2 pandemic is estimated to have resulted in over 500 million infections and over 6 million deaths globally up to May 2022.^1^ A subgroup of COVID-19 survivors report ongoing and debilitating health problems months after mild or severe acute infection,^2^ which cause an increasing burden to society and health care systems.^3^ More information is needed about duration and prevalence and about how vaccination for SARS-CoV-2 affects these symptoms, here referred to as Post COVID-19 condition (PCC).

Commonly reported ongoing symptoms after COVID-19 include fatigue, shortness of breath, myalgia and cognitive problems,^4^ but there is also a wide range of other symptoms involving multiple organ systems.^2^ Lack of a uniform case definition has greatly hampered interpreting prevalence estimates of long-term symptoms after COVID-19. Moreover, estimates vary largely due to different designs, study populations and symptoms assessed.^5-7^ Vaccination for SARS-CoV-2 prior to infection positively affects illness severity in case of break through infections.^8^ Severity of acute disease is associated with development of PCC,^5,9^ and correspondingly it is hypothesised that vaccination prior to infection has a protective effect on developing long-term symptoms due to COVID-19.

The consensus based WHO definition reports that PCC symptoms include, but are not limited to, fatigue, shortness of breath, and cognitive dysfunction, and generally have an impact on everyday functioning.^4^ However, so far few studies on long-term symptoms after COVID-19 include information on impact or severity using validated cut off scores.^10^

Ideally, to define a certain symptom as being part of the set of long-term symptoms related to COVID-19 a causal relation between this symptom and infection with SARS-CoV-22 is assessed. Periods with strict lockdowns and COVID-related measures can affect mood and health in both the infected and uninfected. Therefore, as a first step there is a need for studies that assess to what extend prevalence of fatigue, myalgia and other symptoms after COVID-19 exceed background prevalence in the general population, as well as prevalence in people with acute symptoms who tested negative for SARS-CoV-2. ^11^ Comparing the prevalence of long-term symptoms that present in COVID-19 cases to the prevalence in control groups from the general population as well as from symptomatic test-negative controls can help to identify a set of long-term symptoms that are associated with, or even specific to COVID-19.

In this prospective cohort study we assessed to what extent prevalence and severity of long-term symptoms in mostly non-hospitalized COVID-19 cases exceed the background prevalence and prevalence in people with acute symptoms who tested negative for SARS-CoV-2. Additionally we evaluated the possible protective effect of vaccination prior to infection on long-term symptoms in COVID-19 cases.

## METHODS

### Design, participants and inclusion

Data were collected in the context of the Dutch prospective LongCOVID-study. Details on the study design are described in the study protocol (in preprint).^12^ In this paper we report on COVID-19 cases aged 18 or older three months after a positive SARS-CoV-2 test who joined the study between May 19^th^ 2021 and December 13th 2021. Cases (included within seven days after a positive polymerase chain reaction (PCR) or rapid lateral flow (antigen) SARS-COV-2-test) were recruited from three sources as defined in the study protocol (accepted for publication)^12^ To control for background prevalence of long-term symptoms in individuals without suspected or confirmed COVID-19, two control groups were included. One group consisted of test-negative adults, who were included within seven days after a negative SARS-CoV-2 test and indicated the presence of symptoms as reason to perform the test (test-negative controls); the other control group consisted of adults randomly invited by direct mail from the Dutch population (population controls). Controls with a history of confirmed SARS-CoV-2 infection or a suspected infection that had not been ruled out with a negative test were excluded from the analyses. All participants were asked to voluntarily self-register online on the study’s website (longcovid.rivm.nl) and received questionnaires in their participant portal and reminder e-mails. All participants received a questionnaire at baseline (T0), and a follow-up questionnaire after three months (T3).

### Outcomes

Our primary outcome was the prevalence of having at least one of the significantly elevated symptoms in cases compared to the controls. Secondary outcomes were the severity of the symptoms fatigue, dyspnoea, pain and cognitive impairment assessed with validated cut-off scores, and the difference in prevalence and severity of significantly elevated symptoms between cases that were or were not vaccinated at baseline.

For 41 symptoms participants indicated its presence and for fatigue, pain, cognitive impairment, dyspnoea also symptom severity was assessed. To assess fatigue severity we used the subscale fatigue of the Checklist Individual Strength (CIS);^13,14^ for severity of cognitive impairment the Cognitive Failure Questionnaire (CFQ),^15,16^ for pain severity the bodily pain subscale of the RAND SF-36 Health Status Inventory (SF-36)^17-19^ and for dyspnoea severity the modified Medical Research Council (mMRC) scale.^20^ The mMRC assessed the level of exercise-related dyspnoea in people experiencing breathing difficulties to some extend and was only reported for participants that self-reported to suffer from dyspnoea. Clinical relevant severity of one of the four symptoms was assessed using cut-offs based on previously published norm scores for fatigue (CIS, subscale fatigue, score ≥35),^13,14^ cognitive impairment (CFQ, self-reported cognitive impairment, score ≥44),^15,16^ pain (SF-36, subscale bodily pain, score ≤55),^17-19^ or dyspnoea (mMRC, score ≥1).^20^

At baseline, data was collected on demographics, vaccination status at the moment of a positive SARS-CoV-2 test (cases) or study enrolment (controls), general health status, comorbidities (adapted from Treatment Inventory of Costs in Patients with psychiatric disorders (TiC-P^21^) and on the use of healthcare and medication. Participants were categorized as being *fully vaccinated, partially vaccinated* or *unvaccinated* at baseline (cases and test-negative controls: date of the SARS-CoV-2 test; population controls: date of study enrolment (**supplement**).

### Statistical analyses

Statistical procedures were predefined in a published study protocol (accepted for publication)).^12^ Descriptive statistics were used for participant characteristics, vaccination status and describing acute disease. The prevalence of all symptoms and the severity of fatigue, cognitive impairment, pain and dyspnoea was analysed at T0 and T3, and compared between cases and the two control groups. We considered all of the 41 symptoms reported at T3 in cases and controls. Symptoms with a significant higher prevalence (Benjamini-Hochberg adjusted p-value (p.BH)<0·05) in cases compared to both the test-negative controls and the population controls at T3 were regarded as possibly PCC related. We subsequently report on the primary outcome: prevalence of participants with at least one of the significantly elevated symptoms in cases compared to the controls.

Our primary analysis was based on a complete case analysis, i.e. including only participants that completed both T0 and T3 surveys, without any missing data points as discussed in the study protocol (accepted for publication)).^12^ As a sensitivity analysis, we used four additional scenarios to substitute missing data at T3, and included the symptoms significantly elevated in the primary analysis to assess alternative prevalence of at least one of these symptoms in cases and controls (specified in **supplement**).

To assess the effect of vaccination at baseline, we used the symptoms significantly elevated in cases at T3, and compared the prevalence in fully vaccinated cases versus cases that were partially vaccinated or unvaccinated at the time of their positive SARS-CoV-2 test. A subgroup analysis was performed with cases that were infected when >= 85% of infections was due to the Delta variant in the Netherlands (from July 4^th^ 2021 until December 13^th^ 2021) to preclude impact of different virus variants and other possible bias due to period of inclusion.

For all comparison of prevalence between study groups we used permutation tests stratified for the predefined confounders age, sex, level of education and number of comorbidities (specified in **supplement**) that are possibly associated with the outcomes and might differ between the cases and controls. Subsequently, indirect standardization of symptom prevalence was performed with the cases as reference using the same confounders as for the permutation tests (**supplement**). We identified statistical significance using two-sided 5% significance levels, controlling differences for multiple testing according to the Benjamini-Hochberg procedure,^22^ reporting on adjusted p-values.

Analyses were performed with R version 4.1.0 (packages listed in **supplement**).

### Ethics approval

The Utrecht Medical Ethics Committee (METC) declared in February 2021 that the Medical Research Involving Human Subjects Act (WMO) does not apply to this study as it is survey based (protocol number 21-124/C).

## Role of the funding source

The study is executed by the National Institute for Public Health by order of the Ministry of Health. This means the study is not the result of a competitive grant. The Dutch Ministry of Health, Welfare and Sport does not have a role in the design of this study, its execution, analyses and interpretation of results.

## RESULTS

Until December 13th 2021 14572 participants were enrolled in the study as cases (n=9166), test-negative controls (n=1698) or population controls (n=3708) **(figure 1**). Overall response rate on T3 for these three groups of participants was 71·3% (10389/14572); 72·1% (6614/9166) for the cases, 78·3% (1330/1698) for the test-negative controls and 65·9% (2445/3708) for the population controls.

**Figure 1:**
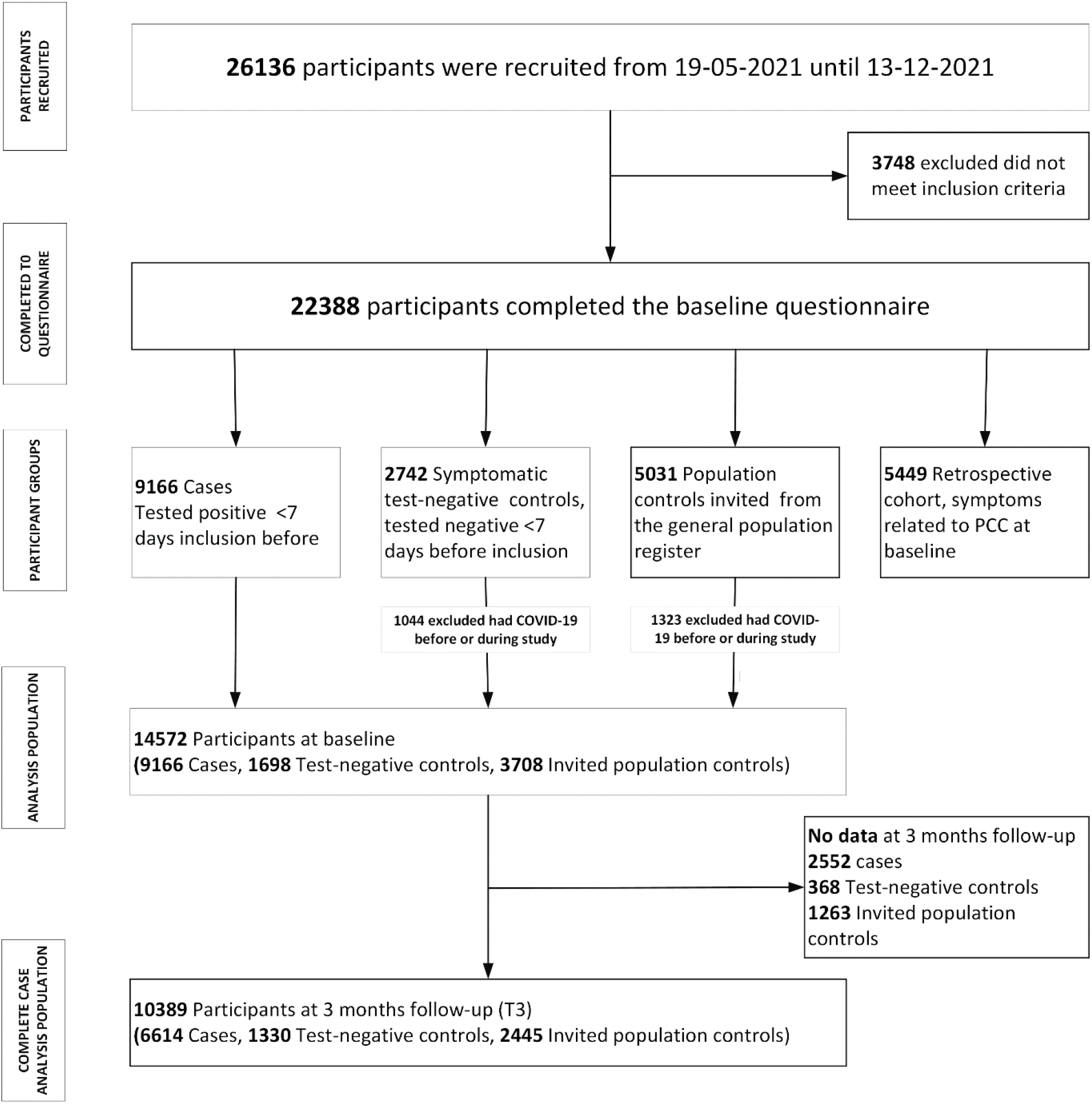
flowchart of study participants and groups included in the study

Baseline characteristics of the cases and controls are shown in **table 1; supplement table S1** shows the characteristics of the complete case study population that completed both T0 and T3 (complete cases).

**Table 1:**
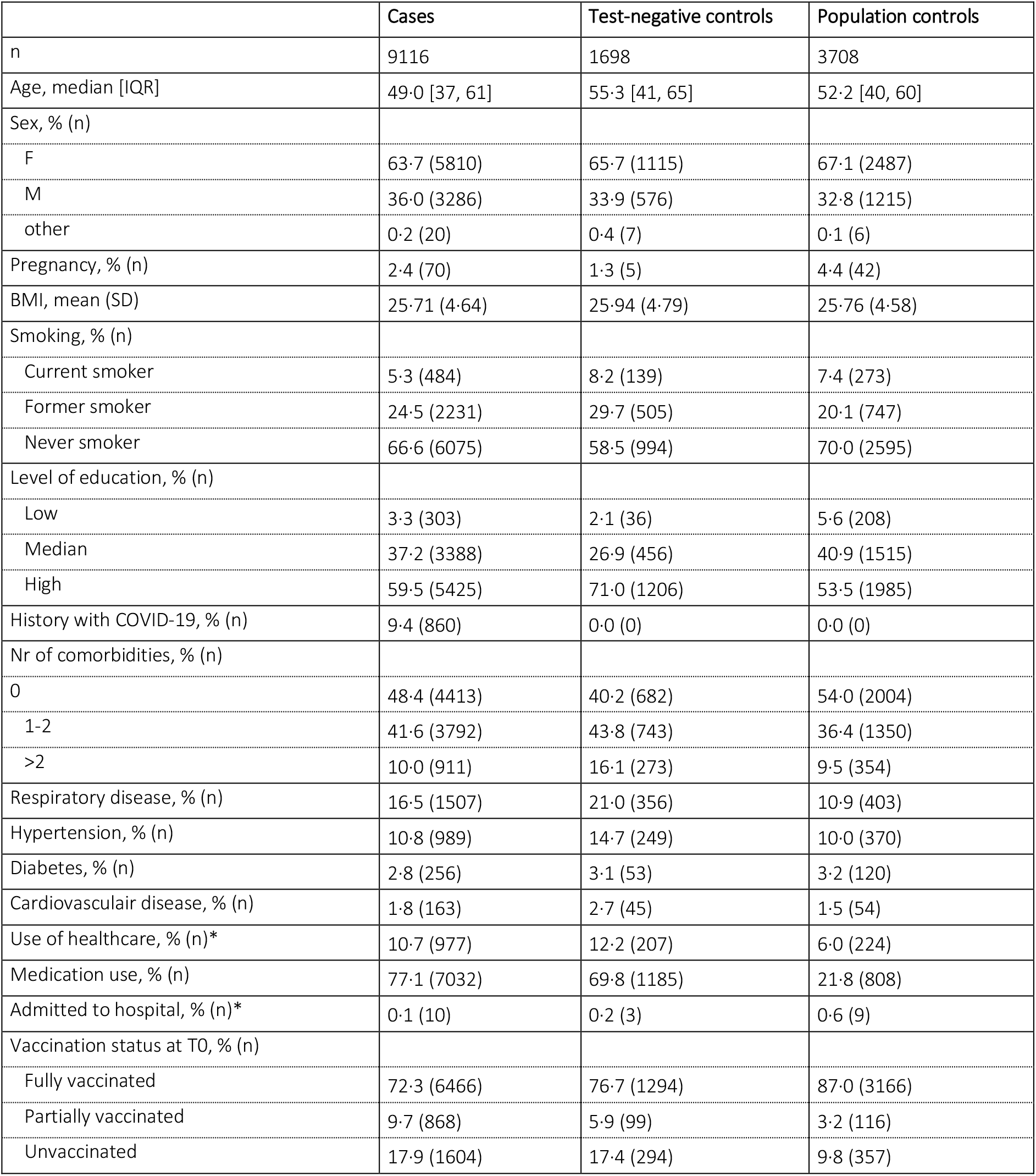
Demographics and acute illness at baseline.

A total of 13 symptoms were significantly elevated in cases compared to both control groups (p.BH<0·05) in the complete case scenario (**figure 2**). The prevalence in cases of fatigue (31·1%), loss of smell (12·0%), dyspnoea (16·4%), difficulty concentrating (15·0%), and difficulties in busy environment (13·1%) showed the largest absolute difference between cases and both control groups. Fatigue (CIS score≥35), cognitive problems (CFQ score≥44) and dyspnoea (mMRC score≥1) at a clinical relevant severity level also had a significantly higher prevalence compared to controls (p.BH<0·05; **table S2 and figure 3**). The subgroup analysis focusing on the Delta variant in the Netherlands (number of cases in complete case analysis for cases n=6440; test-negative controls n=1165, population controls n=2415) did not alter the results (data not shown).

**Figure 2:**
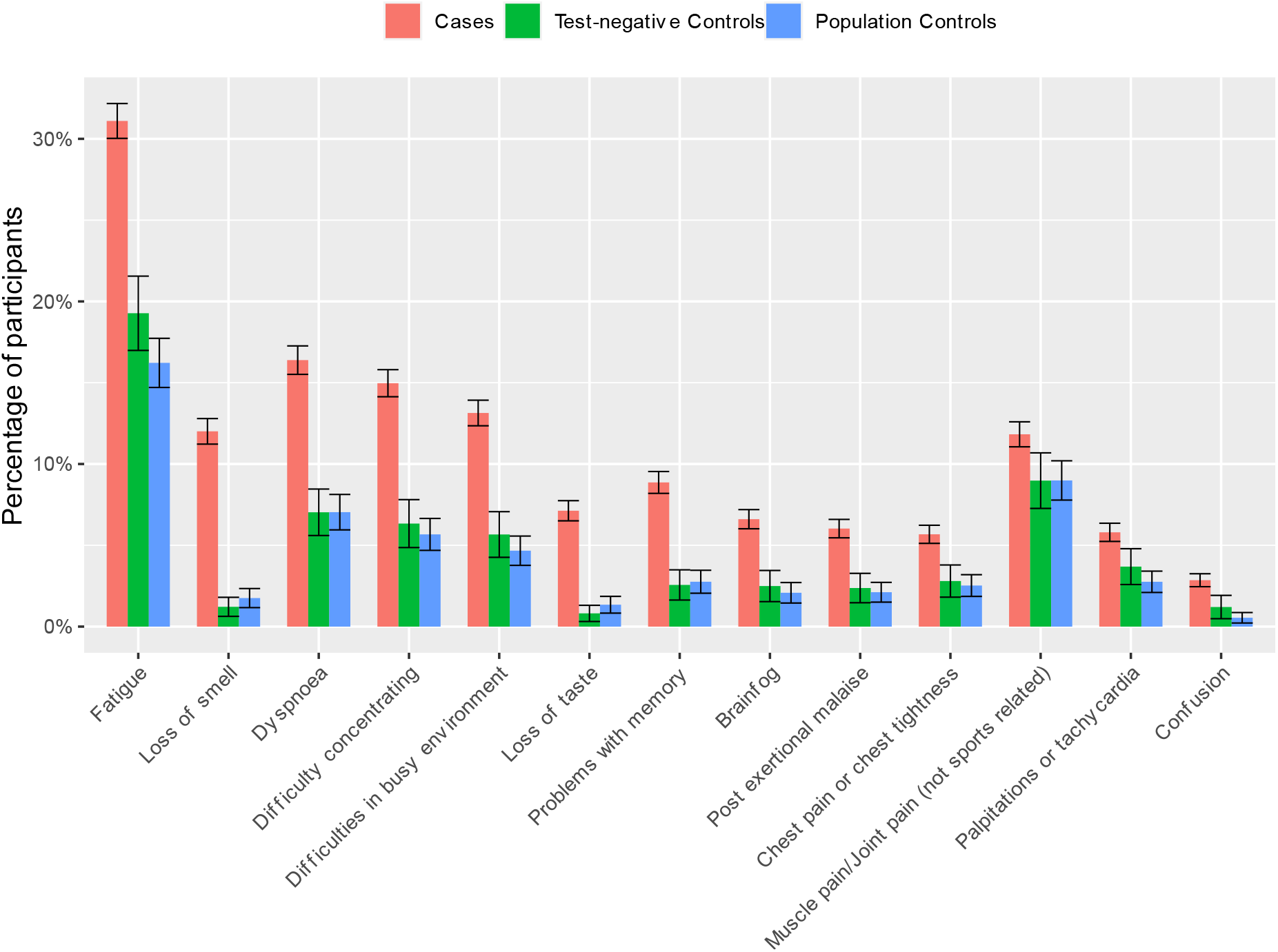
Standardised prevalence (95% confidence intervals) of the 13 symptoms at T3 that were significantly elevated (p.BH<0·05) between cases and both control groups using complete case analysis without substituting for missing values at T3. Symptoms are ranked by the absolute difference in prevalence between cases and test-negative controls.

**Figure 3:**
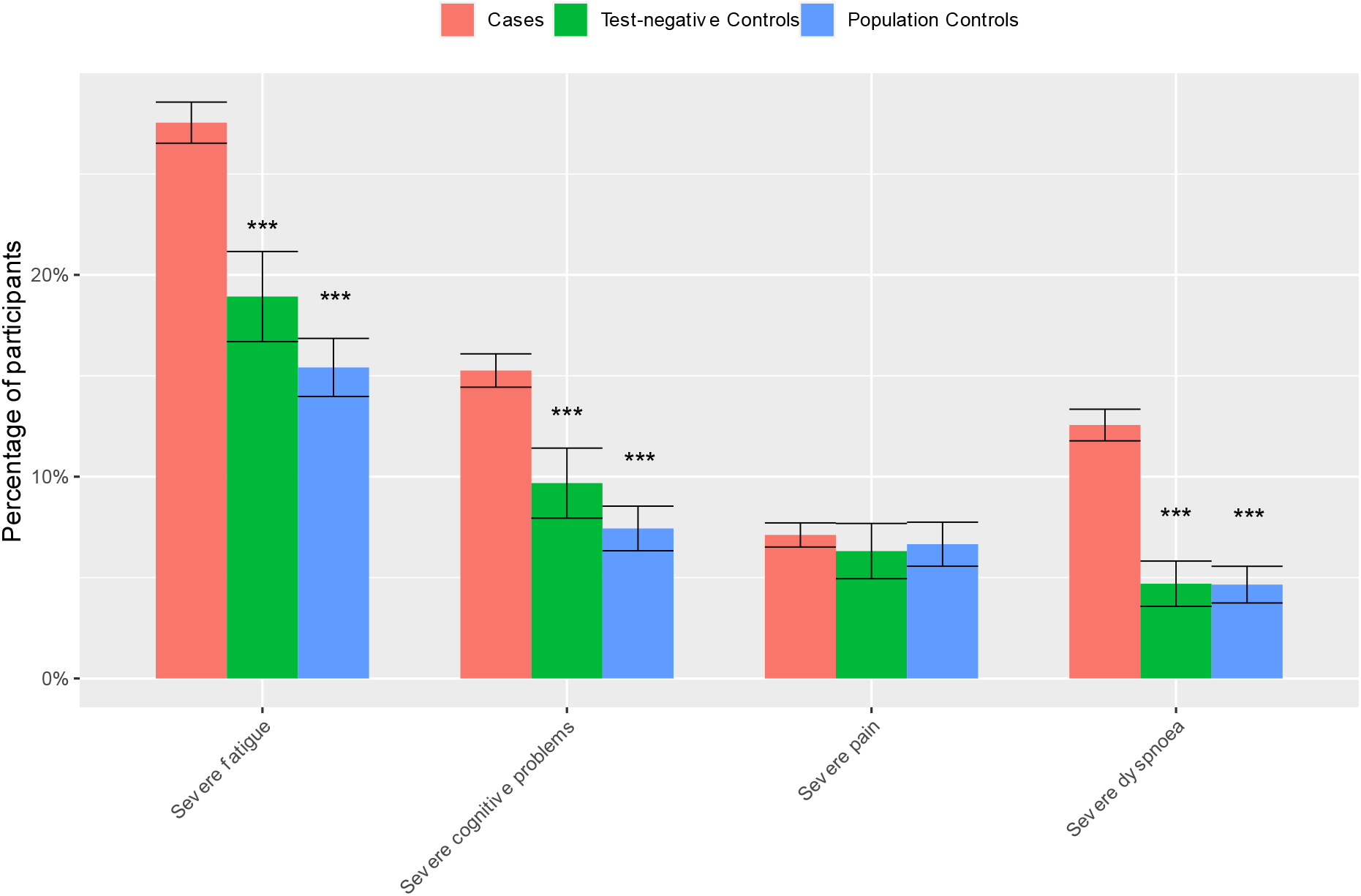
Standardised prevalence (95% confidence intervals) of severity score cut-off values at in cases and both control groups using complete case analysis without substituting for missing values at T3. Severe fatigue: Checklist Individual Strength (CIS), subscale fatigue ≥35, severe cognitive problems: Cognitive Failure Questionnaire (CFQ) ≥44, severe pain: SF-36 subscale bodily pain ≤55, severe dyspnoea: modified Medical Research Council dyspnoea scale mMRC ≥1. ***BH.adjusted p-value <0·001 compared to cases.

Figure 4. shows that in the complete case scenario, almost half (48·5%) of cases reported any of the 13 significantly elevated symptoms., compared to 29·8% of test-negative controls and 26·0% of population controls. The difference between cases and both control groups persisted in the four alternative substitution scenarios, although in the best case scenario the symptom prevalence in both cases and controls was considerably lower (**figure S1**).

**Figure 4:**
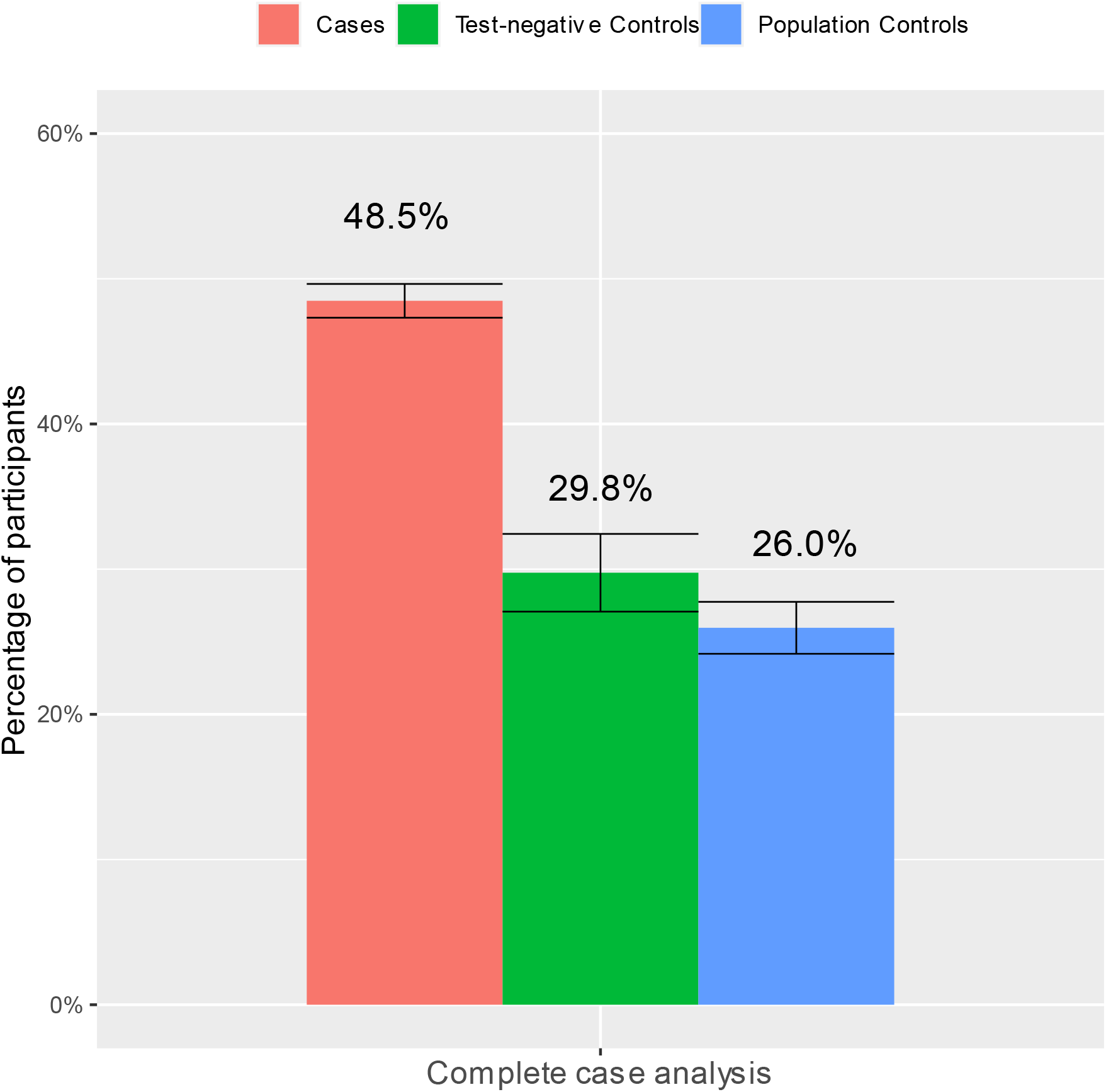
standardized prevalence of at least one of the significantly elevated symptoms at T3 in cases compared to controls in the complete case analysis scenario without substituting for missing values at T3.

Symptoms in cases related to cognition (concentration problems, difficulties in a busy environment, brainfog and confusion, severe cognitive impairment) more often had an onset after baseline, whereas other symptoms, including fatigue, dyspnoea and loss of smell or taste more often persisted from baseline onwards (**figure S2**).

As a result of the timing of recruitment and the vaccination strategy in the Netherlands which initially targeted the older population, we recruited only a small number of unvaccinated cases that were ≥65 years of age, and the analysis on vaccination effect prior to infection could only be performed on cases that were aged <65. **Figure S3** shows that cases <65 years old that were fully vaccinated had a significantly lower prevalence of loss of smell and loss of taste. Other symptoms did not significantly differ between fully, partially or unvaccinated cases in all included cases, and in the subgroup analysis focusing on the Delta variant only (data not shown). Prevalence of at least one of the significantly elevated symptoms in cases was respectively 51·7%, 56·6% and 50·0% in fully vaccinated, partially vaccinated and unvaccinated cases (**figure S4**). Differences in baseline characteristics between fully vaccinated, partially vaccinated and unvaccinated cases, type of vaccination, and effect of vaccination prior to infection on symptoms at T0 are described in the supplement, **table S3 and figure S5**. Among the elevated symptoms, at T0, fatigue, difficulties in a busy environment, chest pain or chest tightness, muscle pain/joint pain and confusion were significantly decreased in fully vaccinated compared to unvaccinated participants (**figure S5**).

## DISCUSSION

In this large observational prospective cohort study, almost half of the COVID-19 cases (48·5%) reported at least one of the possibly PCC-related symptoms three months after SARS-CoV-2 infection, which was approximately 1.5-2 times higher than the background prevalence in the general population (26·0%) and in individuals that likely had another respiratory infection (29·8%). The substantial background prevalence shows that not all reported long-term symptoms after COVID-19 are due to SARS-CoV-2 infection. Only 13 out of 41 considered symptoms were increased in cases compared to controls. Fatigue (31·1%), loss of smell (12·0%), dyspnoea (16·4%), concentration difficulties (15·0%), and difficulties in busy environment (13·1%) showed the largest difference in prevalence between cases and controls. Severe fatigue, severe cognitive impairment and severe dyspnoea were increased in the cases as well. In cases that were fully vaccinated for SARS-CoV-2 prior to infection, the prevalence of loss of smell and loss of taste three months after infection was lower compared to unvaccinated cases (only assessed for the <65 years of age).

### Prevalence of elevated symptoms at three months and defining long-term symptoms that are possibly related to PCC

Our finding that 48·5% of COVID-19 cases had at least one possibly PCC-related symptom three months after the infection is within the range of reported pooled estimates at three months in literature ranging from 29%-55%.^23-25^ However, studies included in pooled estimates are highly heterogeneous with regard to which symptoms studied, the number of symptoms studied, and the included study population. Reported long-term symptoms in literature vary largely due to differences in study design, case definitions and follow-up time. The significantly elevated symptoms in our study are consistent with the WHO definition that claims common PCC symptoms include, but are not limited to, fatigue, shortness of breath and cognitive dysfunction.^4^ In a non-hospitalized population it was reported that fatigue is reported in up to 63% of cases,^5,9,26^ dyspnoea or shortness of breath up to 40%^9,23,27,28^ and cognitive symptoms such as word-finding difficulties, brainfog and concentration problems in up to 40%,^5,23,28^ with a follow-up time ranging from one to 12 months. Part of long-term symptoms after COVID-19 reported in literature, including general malaise, headache, cough and diarrhoea^11,23,29^ were commonly reported in cases in our study, but did not show significant higher prevalence in cases compared to the control groups. These symptoms may therefore well be due to other causes such as other infections or comorbidities. The observed differences between cases and controls indicate that COVID-19 cases are much more likely to report long-term symptoms than people without COVID-19 *and* also much more likely than people with acute symptoms that test negative for COVID. A large part of that last group likely has another respiratory infection.

The impact of COVID-19 thus clearly exceeds the background prevalence, but also the impact of an “average other” respiratory infection. The differences in prevalence between the population controls and test-negative controls indicate that the prevalence of long-term symptoms after other circulating pathogens is much less common compared to symptoms after COVID-19.

### Later onset of cognitive symptoms

Cognitive symptoms such as brainfog, confusion, and concentration difficulties were shown to more often have a later onset after infection compared to other possible PCC symptoms. It remains unclear whether these symptoms are not directly recognised as such by patients in the acute phase, or whether the cognitive impairment becomes apparent or develops only once patients start resuming their usual activities after a period of disease.

### Effect of vaccination

The group of unvaccinated cases was small, and hardly included any people aged >=65. Therefore the vaccination effect was only assessed for cases aged <65. Vaccination had a protective effect only on loss of smell and taste at three months. Vaccination had no effect on the prevalence of other symptoms nor on the overall prevalence of at least one possible PCC-related symptom. Unlike most of the other possible PCC-related symptoms, loss of smell and taste showed a very low background prevalence in controls (below 2%), indicating that the vast majority of these two symptoms in cases is very likely caused by COVID-19. For most other symptoms the background prevalence was substantial and thus in a substantial part of the cases these symptoms are probably not caused by COVID-19. Therefore, for some of these other symptoms our analysis may not have been sensitive enough to address the actual effect of vaccination. In addition, those vaccinated could have risk-factors (for example immune-compromised conditions) which increase the likelihood of a (symptomatic) breakthrough infection as well as the risk for development of long lasting symptoms.^30^ Although, we did control for comorbidities, we cannot exclude the impact of other such risk-factors in our study design. Nevertheless, and although available data on effect of vaccination prior to infection on long-term symptoms is gathered from observational studies, that are also highly heterogenic,^31,32^ the majority of currently available studies do find a decreased prevalence of long-terms symptoms.^31,33-35^

### Representativeness of the study population

The circulating strains during the study period were alfa and delta, but the majority of the cases was included when delta was predominant (>= 85%). Furthermore, our study may have missed participants with no or only minimal acute symptoms at baseline, because they may have been less likely to do a SARS-CoV-2 test. As the severity of acute symptoms is reported to be a risk factor for long-term symptoms after COVID-19,^5,7^ this may have led to an overestimation of prevalence of possible PCC-related symptoms after infection if taken into account all (including asymptomatic) infections. Even though we recruited participants nationwide and from the general population, our study population includes more women, more people with a high level of education, and fewer current smokers than in the overall Dutch population, and may not have been fully representative for the general population in the Netherlands. A striking observation was that at three months after inclusion some symptoms such as fever, coughing and a sore throat were more often reported in controls compared to cases. This may be explained by the fact that a relatively large part of the population controls was recruited around November and December 2021. There was an elevation in influenza cases from half of March 2022 in the Netherlands, possibly resulting in a relatively high background prevalence of these symptoms in controls three months after inclusion.

### Strengths and limitations of the study

A major strength of our study is the fact that we included a large number of both COVID-19 cases as well as two control groups with a prospective follow-up. These two control groups enabled comparison of long-term symptoms in COVID-19 cases with the background prevalence, as well as with the prevalence of symptoms likely due to other respiratory infections. The inclusion of cases and test-negative controls shortly after a positive or negative COVID test and following participants prospectively, is expected to have prevented recall bias with regard to the acute symptoms. Moreover, recruiting from test sites enabled including many COVID-19 cases that were not hospitalised in the acute phase of the disease, and thus more representative for the impact of COVID-19 on the population level.

This study also has limitations. First, data collection only comprised self-reported information through online questionnaires without clinical evaluation of symptoms. However, because of this design we were able to use validated questionnaires with population norm scores, and the aforementioned control groups to correct for the background prevalence. Due to the self-reported results we were not able to preclude possible alternative causes of the reported symptoms. Instead we addressed symptoms that were significantly elevated in cases compared to controls, and report on long-term symptoms. This is in contrast to the WHO definition of PCC which stipulates an exclusion of alternative causes. Moreover, response on the follow-up survey at three months was 71% and it could be that the 29% missing at T3 are to some extent more often the more severely ill, or the other way around more often participants without any symptoms. Therefore, we applied multiple alternative scenarios to substitute the missing values, which showed the robustness of our conclusion based on the complete case analysis. Finally, in our study period two different variants circulated in the Netherlands (alpha and delta). We were not able to look at the impact of the variant on an individual level, but we performed a subgroup analysis on period of inclusion which did not affect results considering prevalence of long-term symptoms in cases or on symptom prevalence in vaccinated compared to unvaccinated cases.

### Conclusions and implications

Three months after infection with SARS-CoV-2 almost half of all COVID-19 cases still experienced at least one symptom which is 1·5-2 times higher prevalence in individuals that likely had another respiratory infection and the background prevalence. Moreover fatigue, cognitive impairment and dyspnoea were more often severe in the cases compared to controls. As these symptoms are significantly elevated in cases compared to controls they are potentially related to PCC. Knowledge on what long-term symptoms are associated with PCC is highly relevant to target possible treatment strategies, for clinical decision-making and for interventional studies to improve long-term outcomes. The substantial background prevalence in the general population further illustrates the challenge in clinical practice to assess for individual patients whether reported long-term symptoms after COVID-19 are due to the SARS-CoV-2 infection, or possible other causes. Vaccination prior to infection in our study population, mostly infected with the delta variant, protected against loss of smell or taste, but not for other long-term symptoms in COVID-19 cases.

## Data Availability

All data produced in the present study are available upon reasonable request to the authors

## DECLARATIONS

## List of abbreviations

CIS: Checklist Individual Strength
CFQ: Cognitive Failure Questionnaire
mMRC: Modified Medical Research Council dyspnoea scale
PASC: Post-Acute Sequelae of SARS-CoV-2 Infection
PCR: polymerase chain reaction
SARS-CoV-2: Severe acute respiratory syndrome coronavirus 2
SF-36: SF-36 item Health Survey
TiC-P: Treatment Inventory of Costs in Patients with psychiatric disorders

## Competing interests

We declare no competing interests.

## Author’s contributions

TM, AH, CW and EF conceptualised the study. EM, CW, KL, HK, AW designed the study protocol and statistical analysis. TM, EM, KL, and JS analysed the data. TM, EM, KL, SB, AT, AH, EF and CW contributed to the data interpretation. TM coordinated the data collection and drafted the manuscript. All authors reviewed and edited revisions of the manuscript, had full access to all the data in the study, and had final responsibility for the decision to submit for publication

## Acknowledgements

We thank Caroline van den Ende for structural updating of literature on PCC.

## Notes

### Competing Interest Statement

The authors have declared no competing interest.

### Funding Statement

The study is executed by the National Institute for Public Health and the Environment by order of the Ministry of Health, Welfare and Sport.

## Reference list

1. World Health Organization. 2021. WHO coronavirus (COVID-19) dashboard. Access date 4 May 2022. [Available from: https://covid19.who.int/.

2. Nalbandian A, Sehgal K, Gupta A, et al. Post-acute COVID-19 syndrome. Nature Medicine. 2021; 27(4): 601–15.

3. Parums DV. Editorial: Long COVID, or Post-COVID Syndrome, and the Global Impact on Health Care. Medical Science Monitor. 2021; 27.

4. Soriano JB, Murthy S, Marshall JC, Relan P, Diaz JV. A clinical case definition of post-COVID-19 condition by a Delphi consensus. Lancet Infect Dis. 2021.

5. Taquet M, Dercon Q, Luciano S, Geddes JR, Husain M, Harrison PJ. Incidence, co-occurrence, and evolution of long-COVID features: A 6-month retrospective cohort study of 273,618 survivors of COVID-19. PLoS Medicine. 2021; 18(9).

6. Zhang X, Wang F, Shen Y, et al. Symptoms and Health Outcomes Among Survivors of COVID-19 Infection 1 Year After Discharge From Hospitals in Wuhan, China. JAMA Netw Open. 2021; 4(9): e2127403.

7. Blomberg B, Mohn KG, Brokstad KA, et al. Long COVID in a prospective cohort of home-isolated patients. Nat Med. 2021; 27(9): 1607–13.

8. Tenforde MW, Self WH, Gaglani M, et al. Effectiveness of mRNA Vaccination in Preventing COVID-19-Associated Invasive Mechanical Ventilation and Death - United States, March 2021-January 2022. MMWR Morb Mortal Wkly Rep. 2022; 71(12): 459–65.

9. Augustin M, Schommers P, Stecher M, et al. Post-COVID syndrome in non-hospitalised patients with COVID-19: a longitudinal prospective cohort study. Lancet Reg Health Eur. 2021; 6: 100122.

10. Malik P, Patel K, Pinto C, et al. Post-acute COVID-19 syndrome (PCS) and health-related quality of life (HRQoL)—A systematic review and meta-analysis. Journal of Medical Virology. 2021.

11. Michelen M, Manoharan L, Elkheir N, et al. Characterising long COVID: a living systematic review. BMJ Glob Health. 2021; 6(9).

12. Mutubuki EN, van der Maaden T, Leung KJ, et al. Prevalence and determinants of persistent symptoms after infection with SARS-CoV-2: Protocol for an observational cohort study (LongCOVID-study). medRxiv preprint. 2022.

13. Worm-Smeitink M, Gielissen M, Bloot L, et al. The assessment of fatigue: Psychometric qualities and norms for the Checklist individual strength. J Psychosom Res. 2017; 98: 40–6.

14. Vercoulen JH, Swanink CM, Fennis JF, Galama JM, van der Meer JW, Bleijenberg G. Dimensional assessment of chronic fatigue syndrome. J Psychosom Res. 1994; 38(5): 383–92.

15. Ponds R, Van Boxtel M, Jolles J. De cognitive failure questionnaire als maat voor subjectief cognitief functioneren. Tijdschrift voor Neuropsychologie. 2006; (2): 37–45.

16. Broadbent DE, Cooper PF, FitzGerald P, Parkes KR. The Cognitive Failures Questionnaire (CFQ) and its correlates. Br J Clin Psychol. 1982; 21(1): 1–16.

17. VanderZee KI, Sanderman R, Heyink JW, de Haes H. Psychometric qualities of the RAND 36-Item Health Survey 1.0: a multidimensional measure of general health status. Int J Behav Med. 1996; 3(2): 104–22.

18. van der Zee KI, Sanderman R. Het meten van de algemene gezondheidstoestand met de RAND-36, een handleiding: umcg /Rijksuniversiteit groningen. Research Institute SHARE. 2012.

19. Aaronson NK, Muller M, Cohen PD, et al. Translation, validation, and norming of the Dutch language version of the SF-36 Health Survey in community and chronic disease populations. J Clin Epidemiol. 1998; 51(11): 1055–68.

20. Mahler DA, Wells CK. Evaluation of clinical methods for rating dyspnea. Chest. 1988; 93(3): 580–6.

21. Bouwmans C, De Jong K, Timman R, et al. Feasibility, reliability and validity of a questionnaire on healthcare consumption and productivity loss in patients with a psychiatric disorder (TiC-P). BMC Health Serv Res. 2013; 13: 217.

22. Hochberg Y, Benjamini Y. More powerful procedures for multiple significance testing. Stat Med. 1990; 9(7): 811–8.

23. Fernández-de-las-Peñas C, Palacios-Ceña D, Gómez-Mayordomo V, et al. Prevalence of post-COVID-19 symptoms in hospitalized and non-hospitalized COVID-19 survivors: A systematic review and meta-analysis. European Journal of Internal Medicine. 2021.

24. Groff D, Sun A, Ssentongo AE, et al. Short-term and Long-term Rates of Postacute Sequelae of SARS-CoV-2 Infection: A Systematic Review. JAMA Netw Open. 2021; 4(10): e2128568.

25. Chen C, Haupert SR, Zimmermann L, Shi X, Fritsche LG, Mukherjee B. Global Prevalence of Post COVID-19 Condition or Long COVID: A Meta-Analysis and Systematic Review. J Infect Dis. 2022.

26. Menges D, Ballouz T, Anagnostopoulos A, et al. Burden of post-COVID-19 syndrome and implications for healthcare service planning: A population-based cohort study. PLoS One. 2021; 16(7): e0254523.

27. Bell ML, Catalfamo CJ, Farland LV, et al. Post-acute sequelae of COVID-19 in a non-hospitalized cohort: Results from the Arizona CoVHORT. PLoS ONE. 2021; 16(8 August).

28. Seeßle J, Waterboer T, Hippchen T, et al. Persistent symptoms in adult patients one year after COVID-19: a prospective cohort study. Clinical infectious diseases : an official publication of the Infectious Diseases Society of America. 2021.

29. Aiyegbusi OL, Hughes SE, Turner G, et al. Symptoms, complications and management of long COVID: a review. J R Soc Med. 2021; 114(9): 428–42.

30. Lipsitch M, Krammer F, Regev-Yochay G, Lustig Y, Balicer RD. SARS-CoV-2 breakthrough infections in vaccinated individuals: measurement, causes and impact. Nat Rev Immunol. 2022; 22(1): 57–65.

31. The effectiveness of vaccination against long COVID. A rapid evidence briefing.: UK Health Security Agency; 2022.

32. Ledford H. Do vaccines protect against long COVID? What the data say. Nature. 2021; 599(7886): 546–8.

33. Antonelli M, Penfold RS, Merino J, et al. Risk factors and disease profile of post-vaccination SARS-CoV-2 infection in UK users of the COVID Symptom Study app: a prospective, community-based, nested, case-control study. Lancet Infect Dis. 2022; 22(1): 43–55.

34. Kuodi P, Gorelik Y, Zayyad H, et al. Association between vaccination status and reported incidence of post-acute COVID-19 symptoms in Israel: a cross-sectional study of patients tested between March 2020 and November 2021. medRxiv preprint. 2022.

35. Al-Aly Z, Bowe B, Xie Y. Long COVID after breakthrough SARS-CoV-2 infection. Nat Med. 2022.

