## supplement for "Prevalence and severity of symptoms 3 months after infection with SARS-CoV-2 compared to test-negative and population controls in the Netherlands"

### Supplementary files

**Table of contents:**


- Supplementary methods p2
- Supplementary results P4
- Table S1: Demographics and acute illness at baseline for complete cases (T0 and T3) P5
- Table S2: Symptoms at three months in cases compared to controls P6
- Figure S1 p9
- Figure S2 p10
- Figure S3 p11
- Figure S4 p12
- Figure S5 p13
- Table S3: Demographics and acute illness at baseline stratified by vaccination status p14
- References p15

#### SUPPLEMENTARY METHODS

*Vaccination status*

Full vaccination at baseline required either one dose of Ad26.COV2.S ‘Janssen’ vaccine at least 28 days prior to a positive test or study enrolment; two doses of BNT162b2 ‘Pfizer/BioNTech’; mRNA-1273 90 ‘Moderna’, or ChAdOx1 ‘Astra Zeneca’ vaccines or a combination of one of these at least 14 days prior to a positive test or study enrolment. Participants were also considered fully vaccinated if they received one vaccination dose at least 14 days prior to a positive test or study enrolment and had had a SARS-CoV-2 infection more than eight weeks before the vaccination. Due to the short time window between date of vaccination and study enrolment in the participants, waning immunity was not taken into account. All other participants that received one or more doses of either vaccine at baseline were regarded *partially vaccinated* – including those that were infected within the 14-day or 28-day immunisation period of the vaccine. Participants were categorized as *unvaccinated* in case no vaccine against SARS-CoV-2 was recorded at baseline.

*Predefined confounders and comparison between groups*

To perform indirect standardization of the primary outcomes with respect to the cases we formed strata using the predefined confounders age (18-45 (young), 46-65 (middle), >65 (old), sex (male and female), educational level (low, median and high) and number of comorbidities (0, 1-2 and >2 comorbidities). For every stratum and group (case and controls) we determined a sample mean and variance, and pooled these to an overall mean and variance of the mean. The overall mean is weighted to the relative size of the stratum (mean) and the square of the relative size (variance of the mean) within the cases. Subsequently, normal approximations from the pooled variances of the mean were used to compute the 95% confidence intervals. We performed permutation tests based on the sum statistic in order to compare symptom prevalence and severity scores between cases and controls, controlling for the same confounders as for the indirect standardization.[1,2] Strata were omitted from testing when containing subjects from one group only, which led to lower sample sizes.

Permutation tests were also used to assess the effect of vaccination before infection on the prevalence of the significantly elevated symptoms in cases, again taking into account age, co-morbidity, sex and education as predefined confounders. Permutation tests were chosen over regression models because of their non-parametric nature: they do not depend on strong assumptions on a functional form, distribution of the response or the error structure to gain full control of type I error. Permutation tests are considered exact tests because they guarantee that the probability of a type I error is exactly equal to the significance level, as chosen by the researcher. They are therefore considered powerful and useful for many situations.[3] Regression models, on the other hand, often have a series of assumptions and can only approximate the probability of type I error. If the assumptions are strongly violated this approximation may be imprecise.

*Scenarios for substitution of missing values at T3*

1. Worst case; including all participants that completed T0, assuming participants that missed T3 had all symptoms reported and all severity scores above cut-off.
2. Best case; including all participants that completed T0, assuming all participants that missed T3 had NO symptoms reported, and NO severity scores above cut-off.
3. Carry forward; including all participants that completed T0, carrying forward the T0 value for all symptoms and severity scores cut-off values to T3.
4. Multiple imputation; multiple imputation was performed with 5 imputations after which data were pooled according to Rubin's rules.

*Multiple imputation*

Multiple imputation was performed with 5 imputations after which data were pooled according to Rubin's rules, using sex, age group, level of education, number of comorbidities and type of participant (case, test-negative control, population control) as predicting variables.

*R packages*

We used Multivariate Imputation by Chained Equations (MICE) package[4] for the multiple imputations and COIN package for the permutation tests [1].

#### SUPPLEMENTARY RESULTS

##### Description of symptoms at T0 and T3

At study enrolment (T0), 98·1% of cases, compared to 99·7% of test-negatives and 42·2% of population controls reported one or more symptoms; respectively 82·9%, 61·8% and 10·2% reported four or more symptoms (**Table S3**). In the test-negative control group, many participants presented with acute symptoms at T0, which to a large extend had decreased at T3. For the population control group symptom prevalence was largely stable between T0 and T3.

##### Selection of symptoms significantly elevated

The selection of the symptoms significantly elevated in cases was solely based on a complete case analysis scenario – not taking into account missing data at three months. In case the alternative scenarios used for substituting missing data would have been used for this selection of symptoms, the larger sample size would likely have resulted in an extended list of symptoms– making it less conservative compared to the complete case scenario.

##### Vaccination status

Of cases, 72·3% (1612) of cases were fully vaccinated at baseline, compared to 76·7% (1294) of test-negatives and 87·0% (3166) of population controls (**table 1**). Respectively, 9·7% (868), 5·9% (99) and 3·2% (116) were partially vaccinated. Differences in vaccination status between cases and controls are explained by timing of recruitment in the population controls and by the vaccination strategy in the Netherlands which initially targeted the older population.

Of cases fully vaccinated, the majority (67·2%) was vaccinated with a Pfizer vaccine, 19·9% with AstraZeneca, 13% with Janssen, 7·5% with Moderna. For the cases that were partially vaccinated, most received a Janssen (51·8%) or Pfizer (38·1%) vaccine (**table S3**). Cases that were fully vaccinated were older (median 51·7 [IQR 40-63] compared to cases that were partially (40·9 [29-49] or unvaccinated (37·8 [27-53]) (**table S3**).

**Table S1: Demographics and acute illness at baseline for complete cases (T0 and T3)**

| | **Cases** | **Test-negative controls** | **Population controls** |
|---|---|---|---|
| N | 6,614 | 1,330 | 2,445 |
| Age, median [IQR] | 51·95 [39·80, 62·59] | 57·37 [44·26, 66·28] | 53·58 [42·65, 60·99] |

| | | | |
|---|---|---|---|
| Sex, % (n) | | | |
| F | 63·3 (4185) | 64·1 (853) | 68·1 (1664) |
| M | 36·5 (2415) | 35·4 (471) | 31·7 (776) |
| Other | 0·2 (14) | 0·4 (6) | 0·2 (5) |
| Pregnancy, % (n) | 2·2 (41) | 0·8 (2) | 4·2 (24) |
| BMI, mean (SD) | 25·73 (4·58) | 25·91 (4·78) | 25·90 (4·66) |
| Smoking, % (n) | | | |
| Current smoker | 4·4 (289) | 7·6 (101) | 5·7 (139) |
| Former smoker | 25·5 (1687) | 29·8 (396) | 21·3 (521) |
| Never smoker | 67·5 (4462) | 59·3 (789) | 71·0 (1735) |
| Level of education, % (n) | | | |
| Low | 3·4 (222) | 2·0 (26) | 5·6 (136) |
| Median | 35·1 (2323) | 25·9 (345) | 38·8 (948) |
| High | 61·5 (4069) | 72·1 (959) | 55·7 (1361) |
| History with COVID-19, % (n) | 9·0 (598) | 0·0 (0) | 0·0 (0) |
| Nr of comorbidities, % (n) | | | |
| 0 | 47·2 (3119) | 40·6 (540) | 54·0 (1320) |
| 1-2 | 42·5 (2811) | 42·6 (566) | 36·5 (893) |
| >2 | 10·3 (684) | 16·8 (224) | 9·5 (232) |
| Respiratory disease, % (n) | 16·6 (1095) | 21·0 (279) | 11·0 (270) |
| Hypertension, % (n) | 12·0 (794) | 15·1 (201) | 11·0 (268) |
| Diabetes, % (n) | 2·9 (192) | 3·6 (48) | 3·5 (86) |
| Cardiovasculair disease, % (n) | 2·0 (130) | 2·8 (37) | 1·4 (35) |
| Use of healthcare, % (n)* | 10·9 (724) | 11·5 (153) | 5·6 (138) |
| Medication use, % (n) | 77·5 (5124) | 68·5 (911) | 22·1 (540) |
| Admitted to hospital, % (n)* | 0·1 (8) | 0·1 (1) | 0·6 (6) |
| Vaccination status at T0, % (n) | | | |
| Fully vaccinated | 75·5 (4902) | 78·8 (1043) | 88·3 (2122) |
| Partially vaccinated | 8·3 (542) | 4·9 (65) | 2·6 (62) |
| Unvaccinated | 16·2 (1049) | 16·3 (216) | 9·1 (219) |

#### Table S2: symptoms at three months in cases compared to controls

| | Complete case analysis (n=) | | | Complete case analysis (n=10389) | | |
|---|---|---|---|---|---|---|
| | **T0** | | | **T3** | | |
| | **Cases, % (95 CI)** | **Test-negative controls, % (95 CI)** | **Population controls % (95 CI)** | **Cases, % (95 CI)** | **Test-negative controls, % (95 CI)** | **Population controls % (95 CI)** |
| Nr of symptoms, median [IQR] | 9.00 [6.00, 13.00] | 6.00 [3.00, 8.00] | 0.00 [0.00, 2.00] | 2.00 [0.00, 4.00] | 1.00 [0.00, 3.00] | 0.00 [0.00, 3.00] |
| Number of symptoms, n (%) | | | | | | |
| 0 | 1.9 (171) | 0.3 (5) | 57.8 (2143) | 35.5 (2348) | 42.6 (566) | 53.8 (1316) |
| 1-2 | 4.8 (442) | 14.4 (245) | 21.4 (794) | 24.8 (1642) | 27.3 (363) | 20.7 (507) |
| 3-4 | 10.3 (942) | 23.5 (399) | 10.5 (391) | 15.4 (1020) | 13.5 (179) | 11.7 (285) |
| >4 | 82.9 (7561) | 61.8 (1049) | 10.2 (380) | 24.3 (1604) | 16.7 (222) | 13.8 (337) |
| Fatigue | **59.9 (58.9- 60.8)** | **41.0*** (38.4- 43.5)** | **13.0*** (11.9- 14.2)** | **31.1 (30.0-32.2)** | **19.3*** (17.0-21.6)** | **16.2*** (14.7-17.7)** |
| Loss of smell | **42.5 (41.5- 43.5)** | **5.7*** (4.4- 7.0)** | **0.8*** (0.4- 1.1)** | **12.0 (11.2-12.8)** | **1.2*** (0.6-1.8)** | **1.8*** (1.2-2.3)** |
| Dyspnoea | **30.2 (29.3- 31.1)** | **19.6*** (17.6- 21.7)** | **3.6*** (3.0- 4.3)** | **16.4 (15.5-17.3)** | **7.0*** (5.6-8.5)** | **7.0*** (5.9-8.1)** |
| Difficulty concentrating | **15.5 (14.7- 16.2)** | **8.2*** (6.7- 9.6)** | **3.8*** (3.2- 4.5)** | **15.0 (14.1-15.8)** | **6.3*** (4.9-7.8)** | **5.7*** (4.7-6.7)** |
| Difficulties in busy environment | **12.4 (11.8- 13.1)** | **7.2*** (5.9- 8.6)** | **4.3*** (3.6- 5.0)** | **13.1 (12.4-13.9)** | **5.7*** (4.3-7.1)** | **4.7*** (3.8-5.6)** |
| Loss of taste | **37.5 (36.5- 38.5)** | **5.4*** (4.1- 6.6)** | **0.5*** (0.2- 0.7)** | **7.1 (6.5-7.7)** | **0.8*** (0.3-1.3)** | **1.3*** (0.8-1.9)** |
| Problems with memory† | **2.4 (2.1- 2.7)** | **1.4*** (0.8- 2.0)** | **1.7* (1.2- 2.1)** | **8.9 (8.2-9.5)** | **2.6*** (1.6-3.5)** | **2.8*** (2.1-3.5)** |
| Brainfog | **6.8 (6.3- 7.4)** | **2.6*** (1.8- 3.5)** | **1.3*** (0.9- 1.7)** | **6.6 (6.0-7.2)** | **2.5*** (1.5-3.5)** | **2.1*** (1.5-2.7)** |
| Post exertional malaise† | **6.9 (6.3- 7.4)** | **2.0*** (1.3- 2.7)** | **0.6*** (0.4- 0.9)** | **6.0 (5.5-6.6)** | **2.4*** (1.5-3.3)** | **2.1*** (1.5-2.7)** |
| Chest pain or chest tightness | **15.3 (14.6- 16.1)** | **5.4*** (4.2- 6.6)** | **1.4*** (1.0- 1.8)** | **5.7 (5.1-6.2)** | **2.8*** (1.8-3.8)** | **2.5*** (1.9-3.2)** |
| Muscle pain/Joint pain (not sports related) | **48.8 (47.8- 49.8)** | **20.8*** (18.6- 23.0)** | **5.4*** (4.7- 6.2)** | **11.8 (11.1-12.6)** | **9.0** (7.3-10.7)** | **9.0*** (7.8-10.2)** |
| Palpitations or tachycardia | **7.1 (6.6- 7.7)** | **4.2*** (3.2- 5.3)** | **2.0*** (1.5- 2.4)** | **5.8 (5.2-6.4)** | **3.7** (2.6-4.8)** | **2.8*** (2.1-3.4)** |
| Confusion | **2.9 (2.6- 3.3)** | **0.8*** (0.4- 1.2)** | **0.6*** (0.3- 0.9)** | **2.9 (2.5-3.3)** | **1.2** (0.5-1.9)** | **0.5*** (0.2-0.9)** |
| Sleep problems | **18.7 (17.9- 19.5)** | **14.3*** (12.5- 16.1)** | **7.3*** (6.4- 8.2)** | 11.8 (11.0-12.6) | 10.3 (8.5-12.0) | 6.1*** (5.1-7.1) |
| Ringing ears | **12.5 (11.9- 13.2)** | **8.4*** (6.9- 9.8)** | **4.8*** (4.1- 5.6)** | 7.7 (7.1-8.3) | 6.2 (4.9-7.5) | 5.7*** (4.7-6.7) |
| Dizziness | **19.6 (18.8- 20.4)** | **8.7*** (7.3- 10.1)** | **2.7*** (2.1- 3.3)** | 6.1 (5.5-6.7) | 4.8 (3.6-6.0) | 3.7*** (2.9-4.6) |
| General malaise | **42.0 (41.0- 43.0)** | **30.0*** (27.5- 32.4)** | **2.6*** (2.1- 3.2)** | 6.1 (5.5-6.7) | 5.0 (3.7-6.3) | 4.4*** (3.6-5.3) |
| Tingling or numbness | **4.4 (4.0- 4.8)** | **2.2*** (1.4- 2.9)** | **1.6*** (1.2- 2.0)** | 4.1 (3.6-4.5) | 3.0 (2.0-3.9) | 1.7*** (1.1-2.2) |
| Headache | **66.1 (65.1- 67.0)** | **42.5*** (39.9- 45.1)** | **14.7*** (13.6- 15.9)** | 20.2 (19.2-21.1) | 19.1 (16.8-21.4) | 17.1*** (15.5-18.6) |
| Shivers | **47.6 (46.5- 48.6)** | **21.5*** (19.3- 23.7)** | **1.8*** (1.4- 2.3)** | 5.2 (4.6-5.7) | 4.3 (3.1-5.6) | 4.4 (3.6-5.3) |
| Mucus from throat or nose | **32.2 (31.3- 33.2)** | **35.7** (33.2- 38.2)** | **5.9*** (5.1- 6.8)** | 6.9 (6.3-7.5) | 6.2 (4.8-7.7) | 6.9 (5.8-8.0) |
| Fever | **44.7 (43.7- 45.7)** | **20.7*** (18.5- 22.9)** | **1.2*** (0.8- 1.5)** | 2.4 (2.0-2.8) | 1.9 (1.1-2.8) | 3.6[##] (2.8-4.4) |
| Earache | **11.1 (10.5- 11.7)** | **7.6*** (6.2- 9.1)** | **1.2*** (0.8- 1.6)** | 2.0 (1.6-2.3) | 1.6 (0.8-2.3) | 1.6 (1.0-2.1) |
| Anxiety† | 1.7 (1.4- 2.0) | 1.0* (0.5- 1.6) | 1.6 (1.1- 2.0) | 2.2 (1.9-2.6) | 1.8 (1.1-2.6) | 1.8 (1.2-2.5) |

| | | | | | | |
|---|---|---|---|---|---|---|
| New onset allergy† | 0.0 (0.0- 0.1) | 0.0 (0.0- 0.0) | 0.1# (0.0- 0.2) | 0.4 (0.3-0.6) | 0.0 (0.0-0.1) | 0.0** (0.0-0.0) |
| Sore eye | 5.9 (5.4- 6.3) | 4.6 (3.5- 5.8) | 1.1*** (0.7- 1.5) | 2.2 (1.9-2.6) | 1.9 (1.1-2.6) | 0.9*** (0.5-1.3) |
| Winter toes | **1.5 (1.3- 1.8)** | **0.6 (0.2- 0.9) ** ** | **0.9** (0.5- 1.2)** | 1.5 (1.2-1.8) | 1.1 (0.5-1.7) | 1.1 (0.6-1.5) |
| Decreased appetite | 29.2 (28.3- 30.2) | 11.5 (9.8- 13.2) | 1.7*** (1.2- 2.1) | 2.9 (2.5-3.3) | 2.5 (1.6-3.5) | 2.1 (1.5-2.8) |
| Menstruation problems† | 1.2 (1.0- 1.5) | 0.9 (0.4- 1.4) | 1.6 (1.2- 2.0) | 2.8 (2.4-3.2) | 2.5 (1.5-3.4) | 1.7** (1.1-2.2) |
| Rash | 2.3 (2.0- 2.6) | 1.8 (1.2- 2.5) | 1.6*** (1.1- 2.0) | 2.4 (2.0-2.7) | 2.3 (1.4-3.2) | 2.4 (1.7-3.1) |
| Nausea | **13.9 (13.2- 14.6)** | **9.7*** (8.1- 11.3)** | **2.6*** (2.0- 3.1)** | 4.1 (3.6-4.5) | 4.0 (2.9-5.1) | 3.3 (2.5-4.1) |
| Depression† | 1.0 (0.8- 1.2) | 1.6 (0.9- 2.2) | 1.8## (1.3- 2.2) | 2.6 (2.2-2.9) | 2.7 (1.7-3.6) | 1.9* (1.3-2.5) |
| Nosebleed | 2.2 (1.9- 2.5) | 2.5 (1.6- 3.3) | 0.8*** (0.5- 1.1) | 1.4 (1.1-1.7) | 1.6 (0.8-2.3) | 1.1 (0.7-1.6) |
| Neuralgia | 1.1 (0.9- 1.3) | 0.5* (0.1- 0.8) | 0.8 (0.5- 1.1) | 1.1 (0.9-1.4) | 1.5 (0.8-2.1) | 1.4 (0.9-2.0) |
| Vomiting | 2.6 (2.3- 2.9) | 1.8 (1.0- 2.6) | 0.6*** (0.3- 0.8) | 0.6 (0.4-0.8) | 1.0 (0.5-1.6) | 0.7 (0.3-1.0) |
| Altered stool | 15.0 (14.2- 15.7) | 8.2 (6.8- 9.6) | 2.1*** (1.6- 2.6) | 4.4 (3.9-4.9) | 5.0 (3.6-6.3) | 3.1* (2.4-3.9) |
| Sore throat | 51.2 (50.2- 52.2) | 70.1### (67.7- 72.4) | 7.7*** (6.8- 8.6) | 9.9 (9.2-10.6) | 10.5 (8.6-12.3) | 11.3# (10.0-12.7) |
| Coughing | **75.7 (74.9- 76.6)** | **64.4*** (61.9- 66.9)** | **10.1*** (9.0- 11.1)** | 10.7 (10.0-11.5) | 12.0 (10.1-13.9) | 13.2## (11.8-14.7) |
| Abdominal pain | **9.6 (9.0- 10.2)** | **7.0*** (5.6- 8.3)** | **3.1*** (2.5- 3.7)** | 3.7 (3.3-4.2) | 5.1 (3.7-6.4) | 2.9# (2.2-3.7) |
| Runny nose | **82.3 (81.5- 83.1)** | **76.1*** (73.9- 78.4)** | **17.3** (16.0- 18.6)** | 20.1 (19.1-21.0) | 21.8 (19.4-24.3) | 18.7 (17.1-20.3) |
| Sneezing | **67.1 (66.1- 68.1)** | **58.7*** (56.0- 61.3)** | **12.0** (10.9- 13.2)** | 12.6 (11.8-13.4) | 17.9 (15.6-20.2)### | 13.6 (12.2-15.1) |
| **Severity cutoff scores** | | | | | | |
| CIS, subscale fatigue, ≥35 | **52.7 (51.7-53.7)** | **31.4 (29.0-33.8)*** ** | **12.8 (11.7-13.9) *** ** | **27.5 (26.5-28.6)** | **18.9*** (16.7-21.2)** | **15.4*** (14.0-16.9)** |
| CFQ, ≥44 | 9.8 (9.2-10.4) | 10.0 (8.4-11.6) | 7.3 (6.4-8.1) *** | **15.3 (14.4-16.1)** | **9.7*** (7.9-11.4)** | **7.4*** (6.3-8.5)** |
| SF-36 subscale bodily pain, ≤55 | 9.8 (9.2-10.4) | 8.3 (6.9-9.7) | 4.8 (4.1-5.5) *** | 7.1 (6.5-7.7) | 6.3 (4.9-7.7) | 6.7 (5.6-7.7) |
| mMRC, ≥1 | **17.2 (16.4-17.9)** | **9.8 (8.3-11.3) *** ** | **2.2 (1.7-2.7) *** ** | **12.6 (11.8-13.3)** | **4.7*** (3.6-5.8)** | **4.6*** (3.7-5.6)** |

Standardised prevalence (95% confidence intervals) of participants with symptoms at T0 and T3 for cases, test-negative controls and population controls using complete analysis without substituting for missing values at T3. Negative values were truncated at 0.

* BH.adjusted p-value <0·05 compared to cases, prevalence higher in cases than in controls

** BH.adjusted p-value <0·01 compared to cases, prevalence higher in cases than in controls

*** BH.adjusted p-value <0·001 compared to cases, prevalence higher in cases than in controls

### BH.adjusted p-value <0·05 compared to cases, prevalence lower in cases than in controls

#### BH.adjusted p-value <0·01 compared to cases, prevalence lower in cases than in controls

##### BH.adjusted p-value <0·001 compared to cases, prevalence lower in cases than in controls

† Data collected on symptom at T0 only for part of the participants
Bold rows: significant difference between cases and both control groups.
CIS: Checklist Individual Strength
CFQ: Cognitive Failure Questionnaire

SF-36: SF-36 item Health Survey
mMRC: Modified Medical Research Council dyspnoea scale

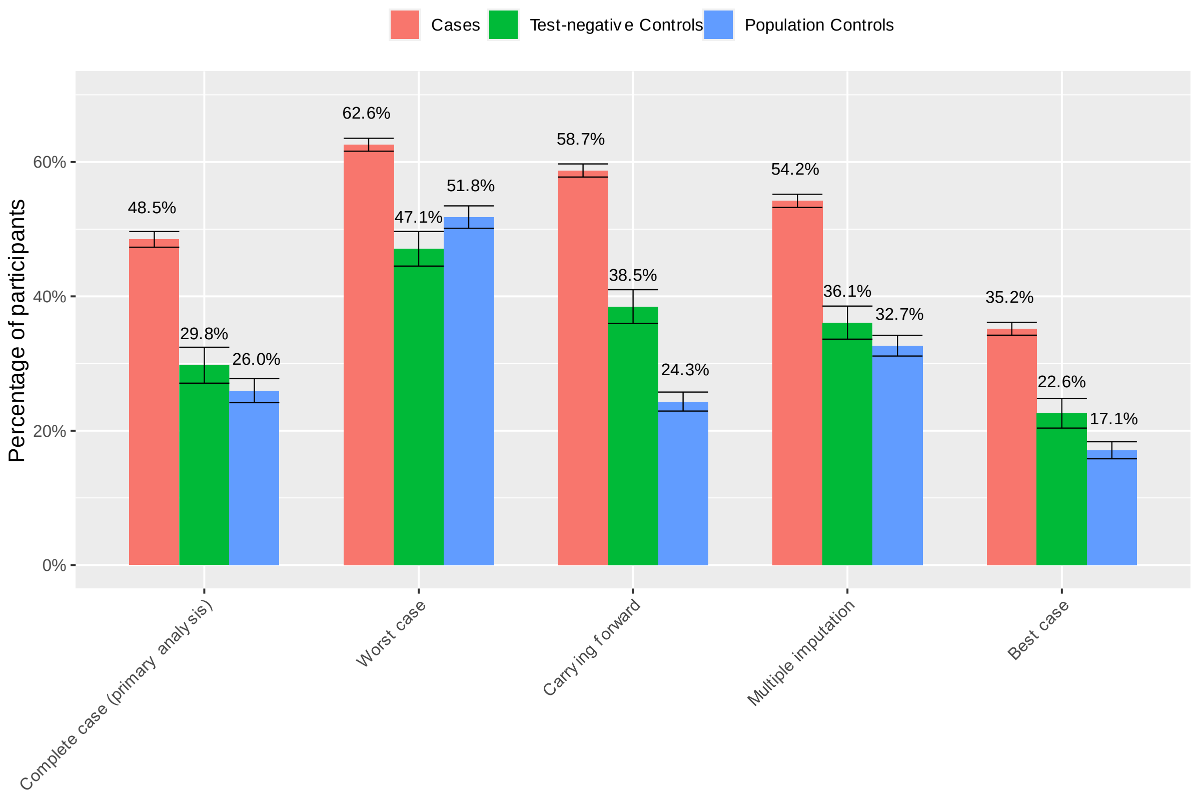


**Figure S1: prevalence of any symptom significantly elevated in cases compared to controls per scenario.** For complete case analysis scenario: n cases: 6614, Test-negative controls: 1330, Population controls: 2445; other scenarios: n cases: 9116, Test-negative controls: 1698 Population controls: 3708. Post exertional malaise and problems with memory were not assessed at T0 and could not be used to predict their prevalence at T3 for the multiple imputation and the carrying forward scenario which may have led to an underestimation of reported prevalence of any symptom significantly elevated in these scenarios.

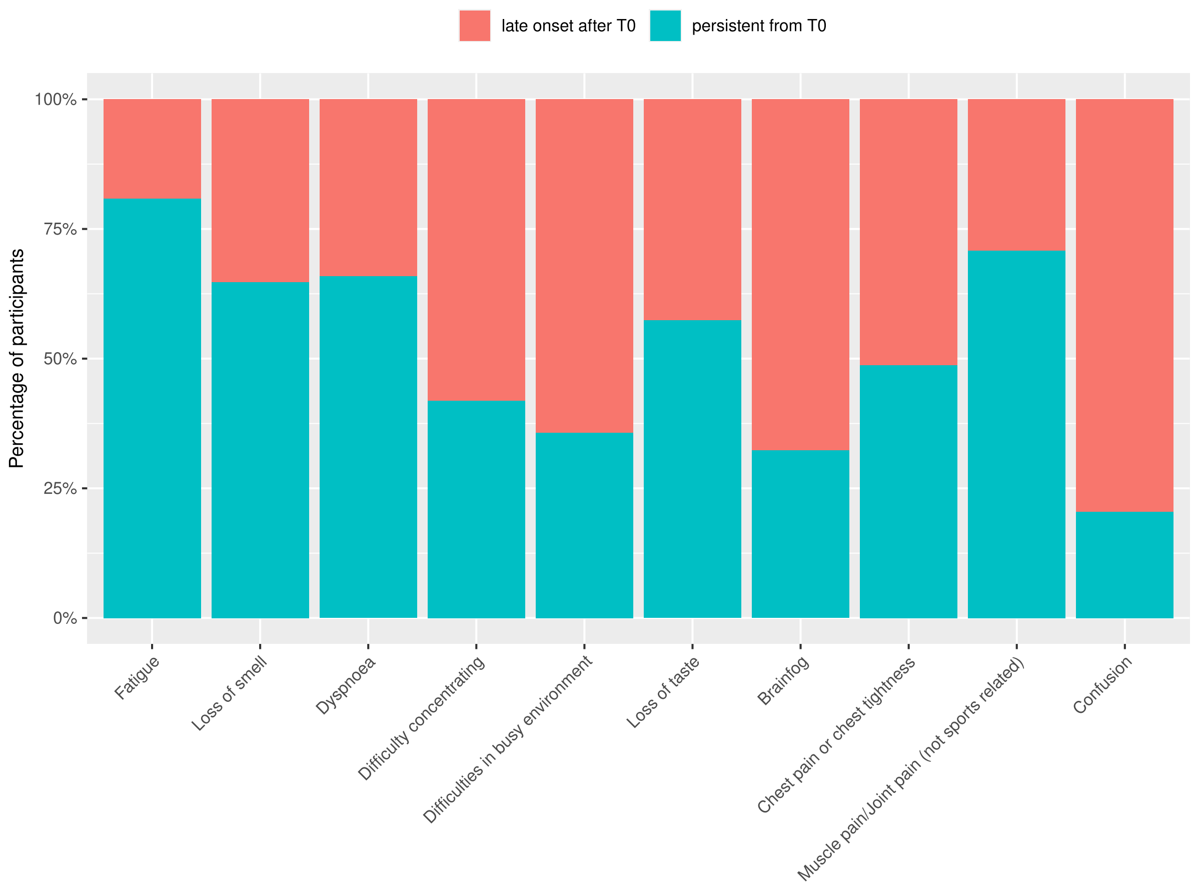


**Figure S2: Development of symptoms that are present at T3, i.e. present at T3 and T0 (persistent from T0 onwards), or present at T3 but not at T0 (late onset after T0) for cases per symptom**. Post exertional malaise and problems with memory were not assessed for all cases at T0 and are omitted from this figure.

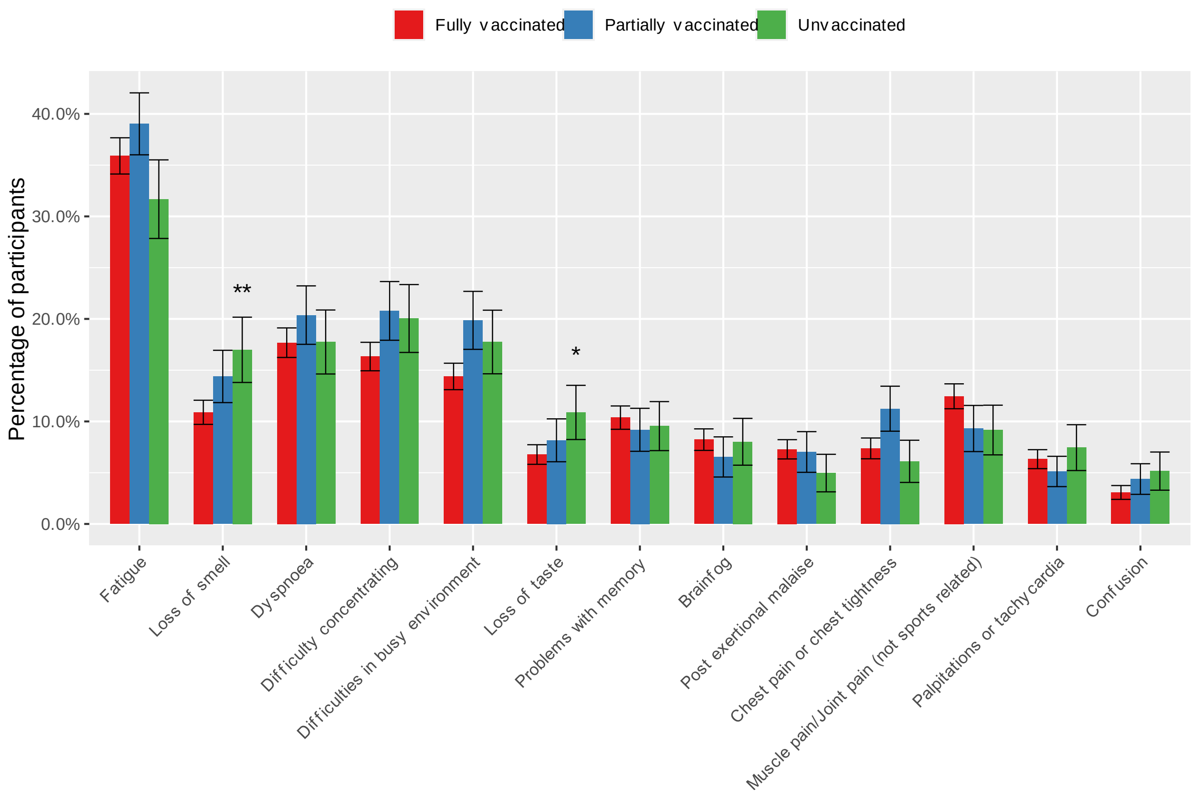


**Figure S3. Standardised prevalence (95% confidence intervals) of participants with symptoms at T3 for cases <65 that were fully, partially or unvaccinated.** Complete analysis without substituting for missing values at T3**.** * BH.adjusted p-value <0·05 compared to vaccinated cases, prevalence higher in unvaccinated cases; ** BH.adjusted p-value <0·01 compared to vaccinated case, prevalence higher in unvaccinated cases

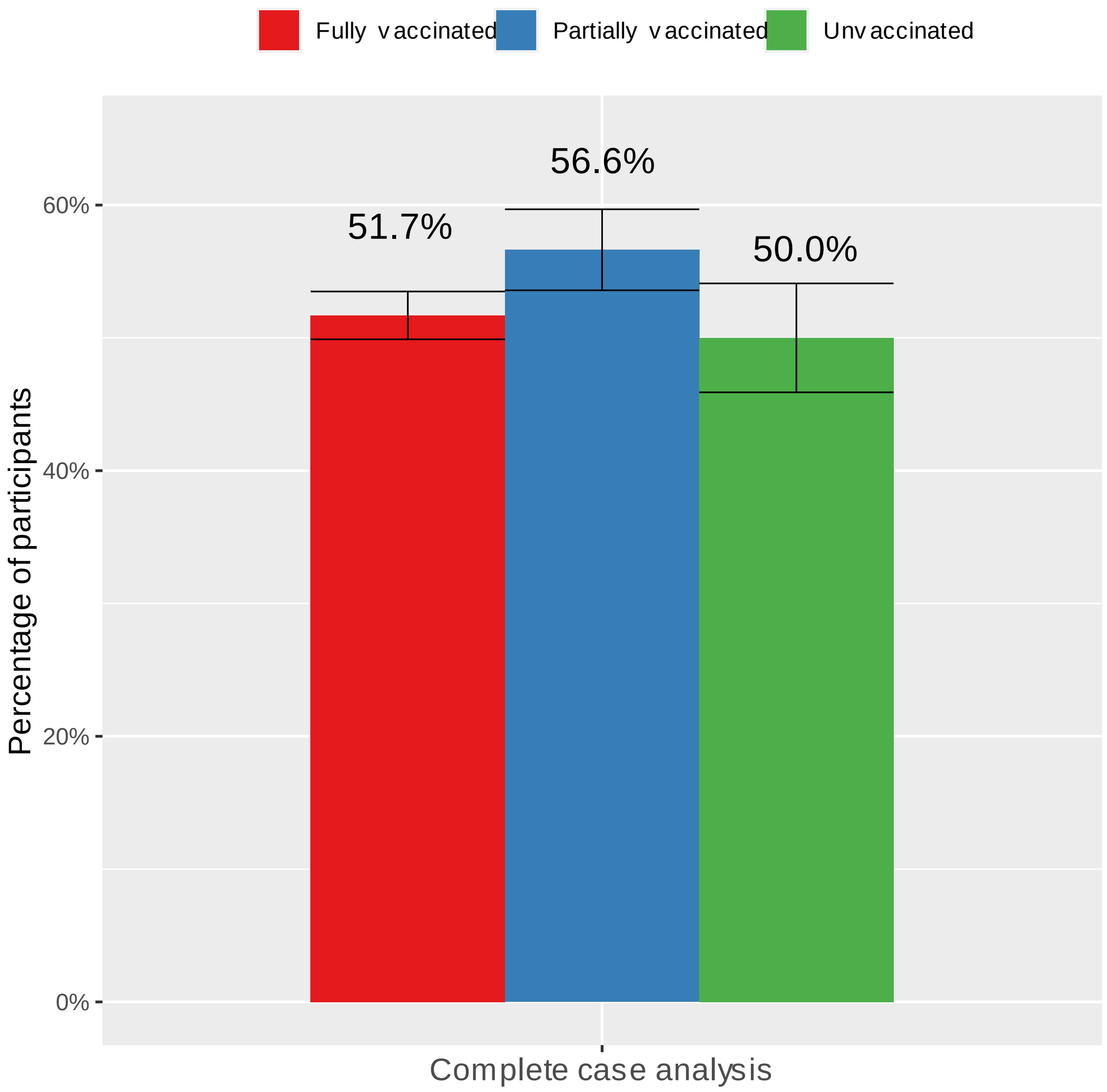


**Figure S4: standardized prevalence of any symptom significantly elevated at T3.** Cases <65 that were fully, partially or unvaccinated using complete analysis without substituting for missing values at T3

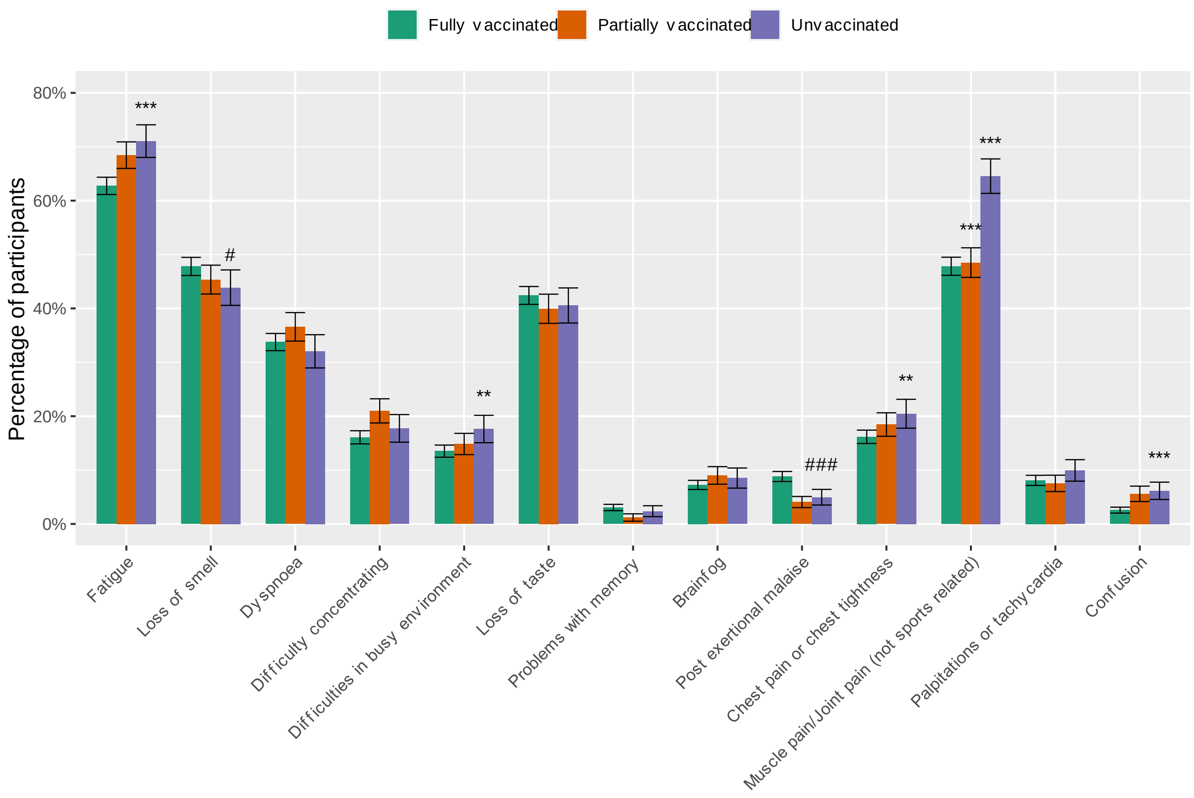


**Figure S5. Standardised prevalence (95% confidence intervals) of participants with symptoms at T0 for cases <65 years old that were fully, partially or unvaccinated.** * BH.adjusted p-value <0·05 compared to vaccinated case, prevalence higher in unvaccinated cases; ** BH.adjusted p-value <0·01 compared to vaccinated case, prevalence higher in unvaccinated cases; *** BH.adjusted p-value <0·001 compared to vaccinated case, prevalence higher in unvaccinated cases; # BH.adjusted p-value <0·05 compared to vaccinated case, prevalence lower in unvaccinated cases; ## BH.adjusted p-value <0·01 compared to vaccinated case, prevalence lower in unvaccinated cases; ### BH.adjusted p-value <0·001 compared to vaccinated case, prevalence lower in unvaccinated cases

#### Table S3: Demographics and acute illness at baseline stratified by vaccination status

| | Fully vaccinated | Partially vaccinated | Unvaccinated |
|---|---|---|---|
| n | 8,058 | 1,771 | 993 |
| **Demographics** | | | |
| Age, median [range] | 51.7 [40, 63] | 40.9 [29, 49]] | 37.8 [27, 53] |
| Sex, % (n) | | | |
| F | 62.6 (5048) | 63.8 (1130) | 71.6 (711) |
| M | 37.2 (2994) | 35.7 (633) | 28.1 (279) |
| other | 0.2 (16) | 0.5 (8) | 0.3 (3) |
| Pregnancy, % (n) | 2.6 (59) | 1.4 (10) | 4.9 (25) |
| BMI, mean (SD) | 26.09 (4.79) | 24.99 (4.40) | 24.88 (4.45) |
| Smoking, % (n) | | | |
| Current smoker | 5.2 (423) | 5.8 (103) | 6.9 (69) |
| Former smoker | 26.2 (2110) | 18.3 (324) | 19.4 (193) |
| Never smoker | 65.7 (5297) | 70.7 (1252) | 68.1 (676) |
| Level of education, % (n) | | | |
| Low | 3.8 (305) | 1.1 (20) | 3.1 (31) |
| Median | 36.7 (2961) | 34.8 (617) | 46.0 (457) |
| High | 59.5 (4792) | 64.0 (1134) | 50.9 (505) |
| History with COVID-19, % (n) | 9.3 (753) | 2.3 (40) | 17.4 (173) |
| Nr of comorbidities, % (n) | | | |
| 0 | 45.1 (3634) | 60.6 (1074) | 57.0 (566) |
| 1-2 | 43.6 (3511) | 33.9 (600) | 34.4 (342) |
| >2 | 11.3 (913) | 5.5 (97) | 8.6 (85) |
| Respiratory disease, % (n)** | 18.0 (1449) | 11.6 (205) | 13.4 (133) |
| Hypertension, % (n) | 12.7 (1020) | 5.4 (96) | 4.8 (48) |
| Diabetes, % (n) | 3.6 (288) | 0.8 (14) | 0.8 (8) |
| Cardiovascular disease, % (n)*** | 2.2 (174) | 0.5 (8) | 0.2 (2) |
| Use of healthcare, % (n)* | 10.1 (815) | 8.9 (158) | 15.5 (154) |
| Medication use, % (n)* | 77.0 (6206) | 76.1 (1348) | 76.9 (764) |
| Admitted to hospital, % (n)* | 0.1 (4) | 0.1 (1) | 0.6 (6) |
| Vaccine type % (freq) | | | |
| AstraZeneca | 19.9 (1601) | 2.3 (41) | NA |
| CureVac | 0.0 (0) | 0.1 (1) | NA |
| Janssen | 0.1 (9) | 51.8 (916) | NA |
| Moderna | 7.5 (606) | 3.4 (60) | NA |
| Pfizer | 67.2 (5413) | 38.1 (674) | NA |
| combination | 5.2 (418) | 4.1 (72) | NA |
| unknown | 0.1 (11) | 0.2 (3) | NA |

**References:**

1. Hothorn TH, K.; van de Wiel, M.A.V.; Zeileis, Al. Implementing a Class of Permutation Tests: The coin Package. *J Stat Softw*. 2008; **28**(8): 1-23.
2. Ferreira JA. Some models and methods for the analysis of observational data. *Statistics Surveys*. 2015; **9**: 106-208.
3. Ernst MD. Permutation Methods: A Basis for Exact Inference. *Statist Sci*. 2004; **19**(4): 676-85.
4. van Buuren SG-O, K. mice: Multivariate Imputation by Chained Equations in R. *Journal of Statistical Software*. 2011; **45**(3): 1-67.
